# Impacts of COVID-19 on sick leave

**DOI:** 10.1101/2021.04.09.21255215

**Authors:** Katrine Skyrud, Kjetil Telle, Kjersti Hernæs, Karin Magnusson

## Abstract

**Aim:** To explore sick leave after COVID-19, by comparing doctor-certified sick leave for up to 6 months after testing for SARS-CoV-2 across employees who tested positive and negative.

**Methods:** In all persons (20-70 year of age) with an employment contract, who were tested for the SARS-CoV-2 in Norway from March 1^st^ to November 1^st^ 2020 (N=740 182 with mean [SD] age 39 [13] years, 44% men), we used a difference-in-difference design to contrast doctor-certified sick leave before and after testing, across employees with negative test and positive test by age and sex groups.

**Results:** Sick leave for those testing positive (N=11 414) remained elevated for up to 2 months after testing when compared to those testing negative (N= 728 768), for men and women aged 20-44 and for men aged 45-70 years (relative increase in sick-leave ∼344-415%, (B_all strata_=0.079, 95% CI=0.076, 0.082). The increase in sick leave was prolonged for women aged 45-70 years only, persisting for up to 4 months after testing positive (relative increase = 35%, B=0.010, 95% CI=0.004-0.035).

**Conclusion:** Sick leave following COVID-19 is elevated for up to two to four months after initial infection, thereafter not elevated compared with employees who tested negative for COVID-19. Women aged 45-70 years tend to have a larger impact of COVID-19 on their work ability than men and younger women.

## Introduction

COVID-19 has in several studies been reported to lead to severe long-term health consequences such as respiratory and circulatory dysfunction among those who were hospitalized for the initial disease. Although mild COVID-19 seems to have few long-term consequences for the use of health care services (1), patient-reported complaints such as fatigue, shortness of breath and musculoskeletal pain are common also after mild initial infection (2, 3). Thus, both severe and mild COVID-19 may be hypothesized to lead to temporary or permanent loss of work ability.

The extent to which work ability is affected for the large number of employees going through COVID-19 is currently unknown. Understanding the impacts of long-covid on the work force is important for informing on the continuation of strict lockdown and disease control measures when those at risk of death to an increasing extent are vaccinated. Because long-term loss of work ability will be far more costly than short contacts with health care, improved understanding of sick leave rates following COVID-19 is important when assessing the societal costs of the pandemic.

Both the prevalence of symptomatic COVID-19 and the prevalence of sick-leave are known to greatly vary with age and sex (4, 5), and it may be hypothesized that women and the elderly are worse off regarding their work ability after having undergone COVID-19, than men and younger persons. Thus, we aimed to explore the impacts of COVID-19 on sick leave for up to 6 months after testing positive or negative for SARS-CoV-2, for all employees in Norway stratified by age and sex.

## Methods

### Design & data sources

We utilized population-wide longitudinal registry data for all employees in Norway to estimate the long-term impacts of COVID-19 on sick leave by applying an observational pre-post design with a comparison group. The BeredtC19-register is an emergency preparedness register. Its main aim is to provide rapid knowledge about the pandemic and its countermeasures. It includes data from the Surveillance System for Communicable Diseases (MSIS) (every polymerase chain reaction tests (PCR) for SARS-CoV-2 in Norway), the Norway Control and Payment of Health Reimbursement (KUHR) Database (all consultations with all general practitioners and emergency primary health care including doctor-certified sick leave), the National Population Register (age, sex, country of birth, date of death), the Norwegian Patient Register (all in- and outpatient hospital contacts), as well as the Employer- and Employee-register. The individual level data are linked over time and across data sources by the unique personal identification number provided every Norwegian resident at birth or upon immigration.

The establishment of an emergency preparedness register forms part of the legally mandated responsibilities of The Norwegian Institute of Public Health (NIPH) during epidemics. Institutional board review was conducted, and The Ethics Committee of South-East Norway confirmed (June 4th 2020, #153204) that external ethical board review was not required.

### Population

Our population included every Norwegian resident on January 1^st^ 2020 who had at least one employment contract of more than 1 weekly working hour, and had been tested for SARS-CoV-2 by a PCR-test from March 1^st^ to November 1^st^ 2020 (non-residents, e.g. tourists, could not be included). We constructed the dataset so that we could follow the employee for at least two months before and at least three months after the test.

### COVID-19

We studied all employees with a PCR-tests for SARS-CoV-2 in Norway, contrasting employees who tested positive with employees who tested negative (using the first available test date).

### Outcomes

Our outcome was doctor-certified (all-cause) sick leave in a given week. We used the reimbursement code L1 as reported from every physician in primary care (general practitioners and emergency wards). All Norwegian employees, both in the public and the private sector, are mandatory members of the national sick leave insurance scheme, which ensures full reimbursement of earnings lost due to sickness up to a generous ceiling and limited to one year. Sickness absence for more than 3 days (private sector), sometimes 7 or more (public sector), needs to be certified by a medical doctor. Because employees who stop working (if e.g. hospitalized with COVID-19 or laid off) do not need physician-certified sick leave, we also studied all-cause health care contacts as outcome (primary, outpatient and inpatient specialist care).

### Statistical analyses

After drawing initial plots of raw data for the entire sample, we used a difference-in-difference approach (6,7) to study the weekly change in sick leave rates from 3 months prior to test week, to 6 months after test week across employees with no COVID-19 (negative test) and with COVID-19 (positive test).

To estimate the impact of COVID-19 on sick leave, we used a standard difference-in-differences regression model with sick leave as the outcome variable (6,7). The main independent variables captured, first, that the individual was in the group that had tested positive (vs. negative), and, second, that the doctor-certified sick leave (dependent variable) occurred in the given period after test week (vs. all 3 months before test week). The interactions between these two sets of independent variables are the classical difference-in-differences estimator, capturing the absolute impact (in percentage points) of COVID-19 on the sick rate. We also calculated the relative impact by dividing the absolute impact estimate by the sick leave rate of those testing negative in the period before test week. We allowed for correlations across multiple observations for the same individual over time (clustered standard errors) and adjusted for the following potential confounders: Comorbidities (categories 0, 1, 2 or 3 or more comorbidities) based on risk conditions for COVID-19 defined by an expert panel (8), birth country (Norway/abroad) and calendar month (12 categories). The differences-in-difference model was run for the following strata of age and sex: Men 20-44 years, women 20-44 years, men 45-70 years and women 45-70 years.

In addition to providing estimates for the impact of COVID-19 on sick leave 0-2 months, 3-4 months and 5-6 months after the test, we also used the model to present plots with rates of sick leave for those with and without COVID-19 by 4-weeks periods before and after the test, by our groups of age and sex. We repeated the analyses using all-cause health care contacts as outcome (primary, outpatient and inpatient specialist care) to explore the probability that our analyses of sick leave were affected by healthy worker selection bias. All analyses were run in STATA MP v.16.

## Results

We studied in total 740 182 employees of which 11 414 (1.54 percent) tested positive. Of these, and among 209 984 men aged 20-44 years and 114 052 men aged 45-70 years, 1.9 percent and 2.0 percent tested positive, respectively. Among 268 259 women aged 20-44 years 1.31 percent tested positive and among 147 887 women aged 45-70 years 1.2 percent tested positive. For all the strata, employees who tested positive were more often born abroad (33 percent) than employees with a negative test (45 percent men and 16 percent born abroad).

Figure 1 shows that employees testing positive had a sick leave of 2.5 percent in the 3 months before testing positive, increasing to 28.5 percent in the test week and dropped to pre-testing levels at month 3 (2.8 percent) and 4 (2.2 percent) after testing. Employees testing negative had 2.0 percent sick leave in the 3 months before testing, increasing to 9.0 percent in the test week, and returning toward pre-testing level in month 3 (2.6 percent) and 4 (2.5 percent) after testing (Figure 1). Possibly related to the pandemic and measures taken to contain it, we thus note that, aside from the couple of months around testing, sick leave remains similar from before testing (2.5 percent) to 5-6 months after testing (2.4 percent) for those who tested positive, but in line with the overall increase in sick leave in Norway during the pandemic (9), it increased somewhat for those who tested negative (2.0 percent to 2.7 percent).

**Figure 1.**
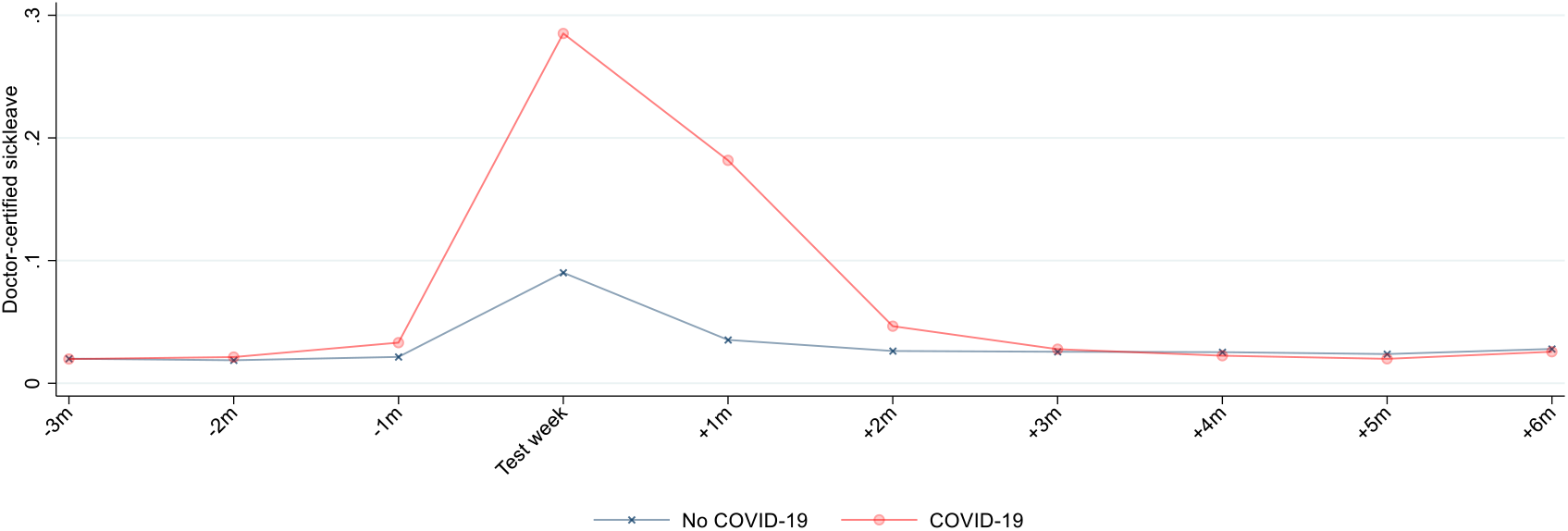
Crude rates of weekly (95% CI) doctor-certified sick leave from 3 months before to 6 months after week of PCR test for SARS-CoV-2, for employees testing negative and positive. +/-1 month includes the 1-4 first weeks after/prior to test week, +/-2 months includes the 5-9 weeks after/prior to test week, etc.

In our differences-in-differences model, we estimated an increase by 7.9 percentage points (see Table 1, upper left cell) in the period 0-2 months after testing positive. Compared with the baseline sick leave rate of 2.0 percent (cf. previous paragraph) for those testing negative in the three months before the testing week, the estimate suggests a relative increase of 392 percent (7.9/2.0). However, at 3-4 months after the test week, the sick leave was similar (−2 percent relative difference) to those who tested negative (Table 1).

**Table 1:** Impact of COVID-19 on doctor-certified sick leave for all employees and subgroups of sex and age. Difference-in-differences estimates for the percentage-points change in the weekly rate of sick leave among employees who tested positive for SARS-CoV-2 from the 3 months before (reference) to the given months after the week of PCR test for SARS-CoV-2, compared with the same change for the employees who tested negative (95% confidence intervals in brackets). Estimates adjusted for comorbidities, birth country and calendar month.

| Months | DID (95% CI) % relative difference |  |  |  |  |  |  |  |  |  |
| --- | --- | --- | --- | --- | --- | --- | --- | --- | --- | --- |
|  | All, 20-70 years |  | Men, 20-44 years |  | Women, 20-44 years |  | Men, 45-70 years |  | Women, 45-70 years |  |
| 0-2 | 0.079 | 392 | 0.053 | 415 | 0.080 | 344 | 0.081 | 477 | 0.132 | 483 |
|  | [0.076,0.082] |  | [0.049,0.056] |  | [0.074,0.085] |  | [0.075,0.088] |  | [0.12,0.14] |  |
| 3-4 | -0.0004 | -2 | -0.005 | -39 | -0.002 | -7 | 0.002 | 11 | 0.010 | 35 |
|  | [-0.002,0.001] |  | [-0.007,-0.003] |  | [-0.005,0.002] |  | [-0.002,0.006] |  | [0.004,0.015] |  |
| 4-6 | -0.006 | -31 | -0.006 | -45 | -0.006 | -26 | -0.005 | -31 | -0.004 | -15 |
|  | [-0.008,-0.005] |  | [-0.008,-0.003] |  | [-0.009,-0.003] |  | [-0.009,-0.002] |  | [-0.009,0.001] |  |

Similar patterns were observed for all the mutually exclusive sex and age groups (Figure 2, Table 1), except for women aged 45-70 years. Their sick leave was elevated 0-2 months after the test (483 percent relative difference) and remained elevated in the months 3-4 after the test (35 percent relative difference) (Figure 2, Table 1). Our analyses of all-cause health care use, in which we explored the potential for healthy worker selection bias showed similar results as our main analyses, implying that our sick leave estimates are not seriously biased by e.g. more non-employment among those testing positive (E-Table 1, E-Figure 1).

**Figure 2.**
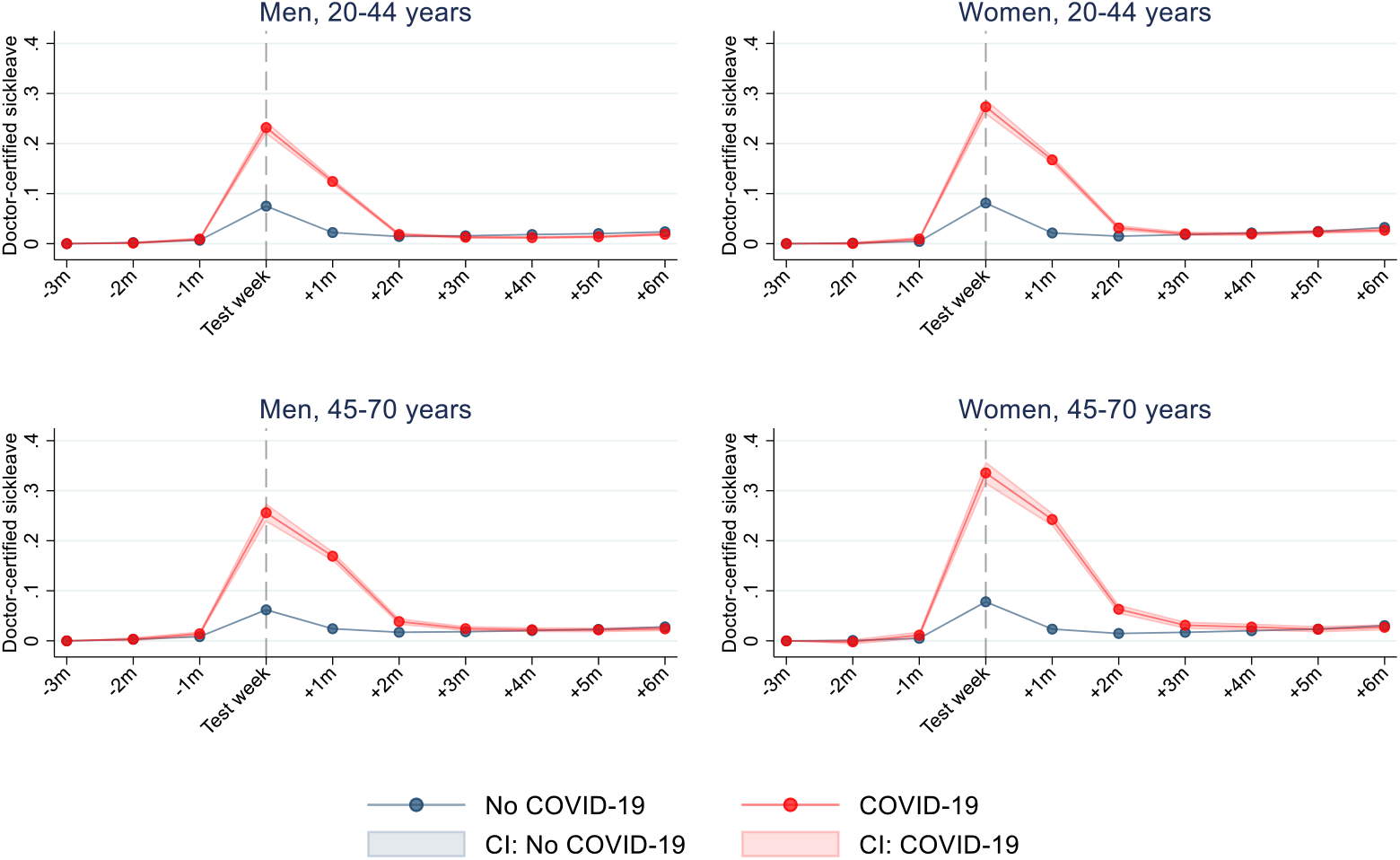
Estimated rates of weekly (95% CI) received doctor-certified sick leave of from 3 months before to 6 months after week of PCR test for SARS-CoV-2, for no COVID-19 and COVID-19 for different sex and age groups. Estimates adjusted for comorbidities, birth country and calendar month. +/-1 month includes the 1-4 first weeks after/prior to test week, +/-2 months includes the 5-9 weeks after/prior to test week, etc.

## Discussion

In all Norwegian employees who were tested for SARS-CoV-2 (N=740 182), we find that the sick leave of employees testing positive was elevated compared to those testing negative for about two months after infection for young and older men. For women, the excessive sick leave of those testing positive compared to those testing negative depended on age, i.e. there was a short-term elevation in sick leave (up to 2 months) for women aged 20-44 years, and a short- and long-term elevation in sick leave (up to 4 months) for women aged 45-70 years.

To our knowledge, this study is the first to explore short- and long-term impacts of COVID-19 on sick leave for employees. Important strengths of our study are the prospective design and use of data that include all employees who were tested for SARS-CoV-2 in an entire country. Another strength is that we could contrast the post-covid sick leave with pre-covid sick leave for the same employee. Thus, by studying the months around the week of testing, and comparing with everyone who tested negative, we could effectively adjust for both time-invariant individual characteristics and seasonal variations in sick leave, transmission and policy measures – including those work-place related - to stop the pandemic.

Whereas several studies have explored the impacts of COVID-19 on long-term health and health care use, especially for patients hospitalized with COVID-19, we are the first to study impacts on sick leave for the vast majority of employees with non-severe initial infection. We believe that sick leave in this group is an important outcome when assessing the total impacts of the pandemic. Though personal and societal costs are large for the patients hospitalized with COVID-19, the sum of these costs may turn out to be modest compared with the overall societal costs of even limited long-term loss of work ability. In our data, for example, of a total of 11 414 employees testing positive for SARS-CoV-2, 544 were hospitalized with COVID-19, and 543 had a minimum of one episode of doctor-diagnosed sick leave with the disease. In this way, the current study sheds new and important light on outcomes that are highly relevant both for the individual health and for society. For example, our findings of a more long-term impact on sick leave among older female employees than among other ages and sex groups, may be important information for employers who are required to facilitate workers in their return to work. The current study shows that adjusting work task or work hours may be particularly important for older female employees who contract COVID-19, to prevent a lengthy sick leave period.

Our study has several important limitations. First, we have no information regarding the length of the sick leave spells. Thus, we cannot study how long an employee is on sick leave, potentially explaining the lower level of sick leave found in our sample than what is reported in official statistics (9). However, doctor-certified sick leave must be renewed regularly, often every fortnight, when symptoms are vague like those typically reported for long-covid (4,10). Moreover, our investigation of potential selection bias due to post-covid non-employment shows that health-care consultations for any cause are affected in a similar way as sick leave (E-Figure 1). This suggests that our sick leave estimates are not seriously biased by e.g. more non-employment among those testing positive. A second limitation may be that we could only include doctor-certified sick leave. In the first 3- or 7-day periods of sick leave, the employee can call in sick without doctor-certification. Thus, we may have underestimated any change in sick leave for short-term spells.

In conclusion, we find that COVID-19 does not elevate sick leave beyond 2 months after testing positive, at least for men aged 20-70 years and for women aged 20-44 years, whereas women aged 45-70 years had elevated sick leave for up to 4 months after testing positive.

## Data Availability

Data are not publicly available

## Acknowledgements

We would like to thank the Norwegian Directorate of Health, in particular Director for Health Registries Olav Isak Sjøflot and his department, for excellent cooperation in establishing the emergency preparedness register. We would also like to thank Gutorm Høgåsen, Ragnhild Tønnessen and Anja Elsrud Schou Lindman for their invaluable efforts in the work on the register. The interpretation and reporting of the data are the sole responsibility of the authors, and no endorsement by the register is intended or should be inferred. We would also like to thank everyone at the Norwegian Institute of Public Health who has been part of the outbreak investigation and response team.

## Author contribution

Kjetil Telle and Karin Magnusson designed the study. Katrine Damgaard Skyrud and Kjersti Hernæs had access to all of the data in the study and takes full responsibility for the integrity of the data and the accuracy of the data analysis. Katrine Damgaard Skyrud and Kjersti Hernæs performed the statistical analyses. Karin Magnusson drafted the manuscript. Kjetil Telle critically evaluated all stages of the research process. All authors contributed with acquisition of data, conceptual design, analyses and interpretation of results. All authors contributed in drafting the article or critically revising it for important intellectual content. All authors gave final approval for the version to be submitted.

## Supplementary file for the paper

**E-Table 1:** Impact of COVID-19 on health care utilization for all employees. Difference-in-difference estimates for the change in the rate of utilizing all-cause health care service per week, after the week of PCR test for SARS-CoV-2.

| Months | DID [95 % CI]. % *Relative difference* |  |
| --- | --- | --- |
| 0-2 | 0.111 [0.11,0.11] | 146 |
| 3-4 | -0.002 [-0.005,0.002] | -2 |
| 5-6 | -0.015 [-0.020,-0.001] | -19 |

**E-Figure 1.**
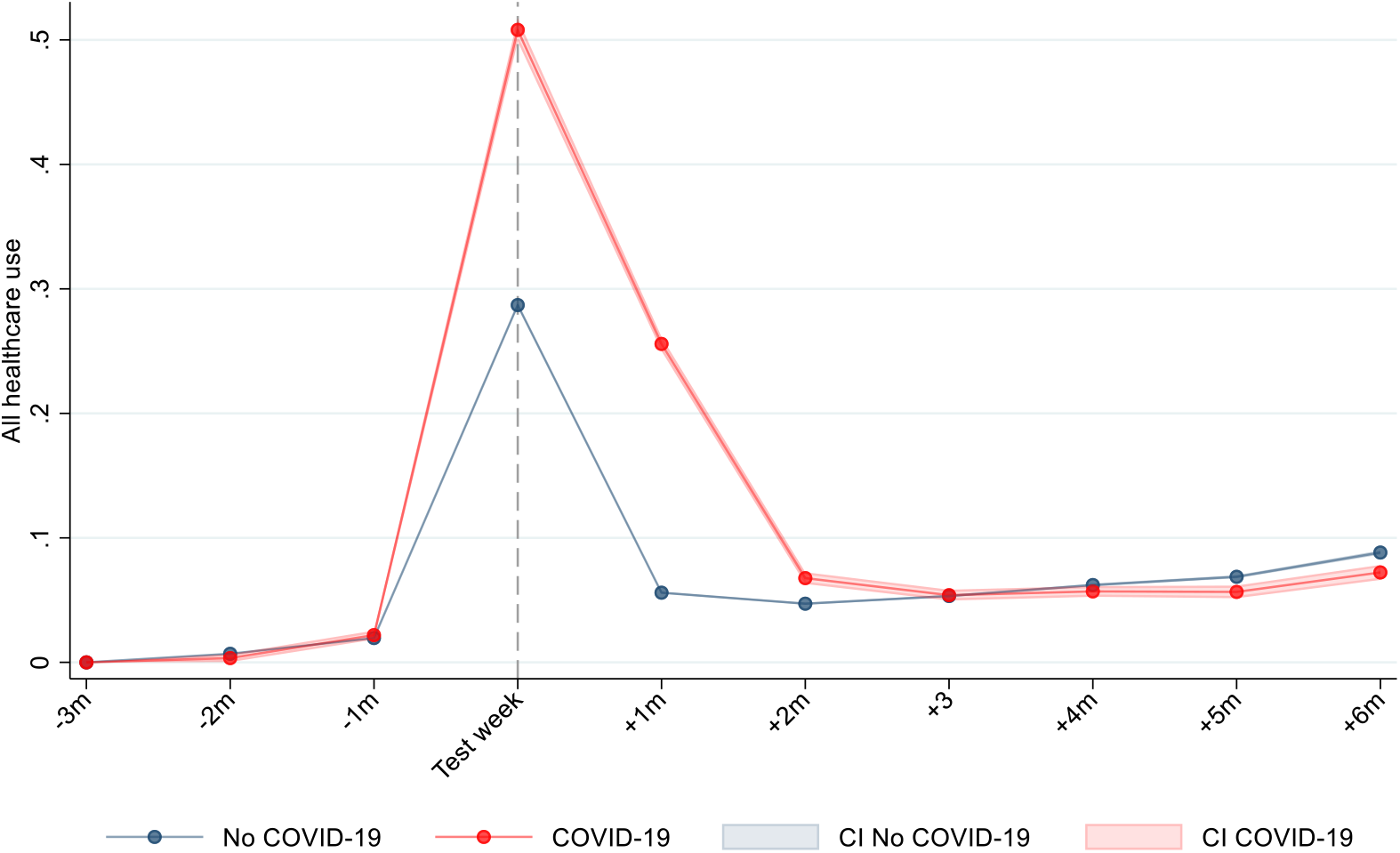
Estimated rates of weekly utilization (95% CI) of any health care (primary and specialist care), from 3 months before to 6 months after week of PCR test for SARS-CoV-2, for no COVID-19 and COVID-19. Estimates adjusted for age, sex, comorbidities, birth country and calendar month. +/-1 month includes the 1-4 first weeks after/prior to test week, +/-2 months includes the 5-9 weeks after/prior to test week, etc.

